# Systematic development and feasibility testing of a multibehavioural digital prehabilitation intervention for patients approaching major surgery (iPREPWELL): A study protocol

**DOI:** 10.1101/2022.10.21.22281380

**Authors:** J Durrand, R Livingston, G Tew, C Gillis, D Yates, J Gray, CJ Greaves, J Moore, A F O’Doherty, P Doherty, G Danjoux, L Avery

## Abstract

Improving outcomes for people undergoing major surgery, specifically reducing perioperative morbidity and mortality remains a global health challenge. Prehabilitation involves the active preparation of patients prior to surgery, including support to tackle risk behaviours that mediate and undermine physical and mental health and wellbeing. The majority of prehabilitation interventions are delivered in person, however many patients express a preference for remotely-delivered interventions that provide them with tailored support and the flexibility. Digital prehabilitation interventions offer scalability and have the potential to benefit perioperative healthcare systems, however there is a lack of robustly developed and evaluated digital programmes for use in routine clinical care. We aim to systematically develop and test the feasibility of an evidence and theory-informed multibehavioural digital prehabilitation intervention ‘iPREPWELL’ designed to prepare patients for major surgery. The intervention will be developed with reference to the Behaviour Change Wheel, COM-B model, and the Theoretical Domains Framework. Codesign methodology will be used to develop a patient intervention and accompanying training intervention for healthcare professionals. Training will be designed to enable healthcare professionals to promote, support and facilitate delivery of the intervention as part of routine clinical care. Patients preparing for major surgery and healthcare professionals involved with their clinical care from two UK National Health Service centres will be recruited to stage 1 (systematic development) and stage 2 (feasibility testing of the intervention). Participants recruited at stage 1 will be asked to complete a COM-B questionnaire and to take part in a qualitative interview study and co-design workshops. Participants recruited at stage 2 (up to twenty healthcare professionals and forty participants) will be asked to take part in a single group intervention study where the primary outcomes will include feasibility, acceptability, and fidelity of intervention delivery, receipt, and enactment. Healthcare professionals will be trained to promote and support use of the intervention by patients, and the training intervention will be evaluated qualitatively and quantitatively. The multifaceted and systematically developed intervention will be the first of its kind and will provide a foundation for further refinement prior to formal efficacy testing.

## Introduction

Approximately 310 million people undergo surgery globally each year [1], and requirement for surgical intervention continues to grow. Improving perioperative outcomes is an ongoing healthcare challenge. In the UK 2.4 million major surgical procedures are undertaken by the National Health Service (NHS) annually [2], with associated perioperative mortality and major morbidity rates estimated at 3.5-4% [3,4] and 15-40% respectively [5]. A single major complication such as wound infection, postoperative pneumonia, myocardial infarction or acute kidney injury profoundly disrupts a patients’ recovery and has major implications for healthcare utilisation. For example, length of hospital stay is increased up to 3-fold [6], risk of re-admission is significantly increased [7], and patients are less likely to be discharged to their home environment [8]. In the longer-term, functional status and quality of life of patients is undermined for several months following discharge, with many individuals never regaining their former independence [9].

Physically and mentally preparing patients for major surgery is one strategy to improving outcomes, a concept known as prehabilitation [10]. Patients with better physical [11], nutritional [12] and mental health [13] encounter fewer complications, leave hospital sooner and experience a faster and more complete recovery, with better preservation of their preoperative independence and quality of life [10]. Optimising the preoperative physical and mental health of individuals in this way carries considerable importance. Co-morbid disease and health risk behaviours render the body less able to tolerate the physiological demand of surgery, thereby elevating the risk of perioperative complications 2-3 fold [10]. Furthermore, anxiety and low self-esteem are also very common in preoperative patients and have shown to increase perioperative risk [13]. These risk factors frequently cluster in surgical patients with at least two evident in 40% of patients presenting for major surgery [14].

Fortunately, scheduled surgery presents a key ‘teachable moment’ to facilitate behavioural change [14]. Patients have been shown to be amenable to optimising their health using behavioural change interventions preoperatively. Furthermore, changes in health behaviours that can increase resilience for surgery and reduce perioperative risks are achievable within 4 weeks [15]. The main pillars of prehabilitation are physical activity and exercise, nutritional optimisation, and support for mental wellbeing [16]. However, interventions to promote smoking cessation [17], alcohol reduction [18] and improved sleep quality [19] may be equally important and should be incorporated into multibehavioural interventions to optimise patient health in the limited time available preoperatively.

Access to preoperative support is a clear patient priority. Prior work has emphasised the importance of improved postoperative functional outcomes from the patient perspective [20], the area of strongest evidence for the benefits of support [15]. At a system level, prehabilitation is now a key recommendation of several national initiatives to improve the quality of UK perioperative care [21, 22]. This shift in focus across perioperative services is now cross-specialty, underlined by the recent reframing of ‘waiting lists’ to ‘preparation lists’ driven in part by the severe impact of the Covid-19 pandemic on surgical waiting times and population health [22].

The Covid-19 pandemic has greatly influenced the delivery of prehabilitation services in the UK over the last 2 years. Several established services that were previously delivering face-to-face interventions were forced to rapidly innovate to deliver remote support to patients. This was at a time when evidence-based remote solutions, including digital interventions, were lacking. The subsequent ‘explosion’ of interest in digital healthcare initiatives has gone some way to help meet this shortfall with NHS organisations often working in partnership with industry to rapidly create solutions. However, the lack of evidence-informed, systematically developed interventions raises questions about effectiveness, replicability, and, of critical importance, uptake and continued engagement by patients and healthcare professionals (HCPs). Uptake and engagement was the topic of an editorial [23] that highlighted the need to address several key questions in the context of intervention development. These include determining whether a digital solution is wanted by patients and HCPs and why; to what extent they believe it would be beneficial; how a digital intervention could be used to optimise outcomes; whether it would be cost-effective; and whether is there a risk of increasing inequalities in perioperative care.

The experience of face-to-face prehabilitation services pre-pandemic indicated that up to 50% of patients were unable or unwilling to engage with this model [24] Barriers include: The requirement to travel, associated cost, inflexibility in terms of time and location, and discomfort in group settings. Digital solutions offer a potential alternative and have been successfully delivered elsewhere in the context of type 2 diabetes management [25] and cardiac rehabilitation [26], and these interventions have observed high levels of patient engagement and health behaviour changes comparable to face-to-face programmes. Given the similarities between these populations and those preparing for major surgery, in terms of age, comorbidity and health behaviour characteristics, it is reasonable to assume that uptake and engagement with a digital intervention preoperatively would be comparable.

The use of digital prehabilitation interventions aligns with wider NHS drivers to incorporate digital technology into patient care [27]. The need for scalability and efficient use of staff time makes digital solutions a logical way forward and has the potential to enhance healthcare systems and service delivery.

In the context of digital health behaviour change, ‘digital exclusion’ is a key concern and has the potential to widen existing health inequalities [28]. Those in the most deprived socioeconomic groups exhibit the highest rates of health risk behaviours that elevate perioperative risk, yet they also face barriers to using digital interventions including access to a device and continued internet access [28]. In addition, the mean age of patients undergoing major surgery is 67 years. Whilst information technology confidence and internet usage in older age groups continues to grow, and a greater proportion of older adults have become familiar with remote services due to Covid, there are still a proportion of this population who are not confident to use digital interventions. As such, utilisation of co-design methods is key to mitigating these inequalities and optimising engagement of patients and HCPs [29]. As with all health behaviour change interventions, prehabilitation interventions are likely to be significantly enhanced by employing a systematic, theory and evidence-informed developmental process in collaboration with stakeholders to increase uptake, engagement, adherence, and overall impact [29].

### Study Aims and objectives

The aim of this study is to systematically develop, and feasibility test a multibehavioural digital prehabilitation intervention for patients approaching major surgery. More specifically, the main objectives are as follows:

1. To develop a theory and evidence-informed digital prehabilitation intervention to target changes in lifestyle behaviours including physical activity, exercise, nutrition, alcohol consumption, sleep, smoking and psychological wellbeing prior to major surgery
2. To develop a theory and evidence-informed training resource for HCPs to promote and support delivery of the digital intervention
3. To assess the feasibility, acceptability, and fidelity of the digital intervention for patients approaching major surgery and supporting HCPs
4. To assess adherence to and completion of the intervention (i.e., do participants work through all components of the intervention and engage with the HCPs providing support?)
5. To assess the feasibility, acceptability, and fidelity of delivery and receipt of the training intervention for HCPs
6. To conduct a qualitative process evaluation with participants (patient and HCPs) to identify determinants of uptake, engagement, continued use and completion of the intervention.
7. To develop a set of implementation strategies with stakeholders to facilitate future implementation of the intervention should it demonstrate to be acceptable and feasible.

Additional objectives are to generate estimates of variability for behavioural outcomes (e.g., physical activity) and outcomes (e.g., quality of life) to inform a sample size calculation for a randomised controlled trial (should the intervention demonstrate acceptability and feasibility), and to undertake a preliminary cost evaluation of the intervention.

## Materials and Methods

### Study setting and design

This two-stage study will be conducted at two NHS Hospital Trusts: South Tees Hospitals NHS Foundation Trust, Middlesbrough, UK and York and Scarborough Teaching Hospitals NHS Foundation Trust, York, UK.

Stage 1 of the study involves the systematic development of an evidence and theory-informed multifaceted behavioural intervention, and stage 2 involves testing the feasibility of the intervention in practice. <u>Figure 1 presents a SPIRIT schedule of enrolment, intervention and assessments for study stage 2 and an overview of stage 1 and 2 design and timelines is presented in figure 2.</u>

**Figure 1:**
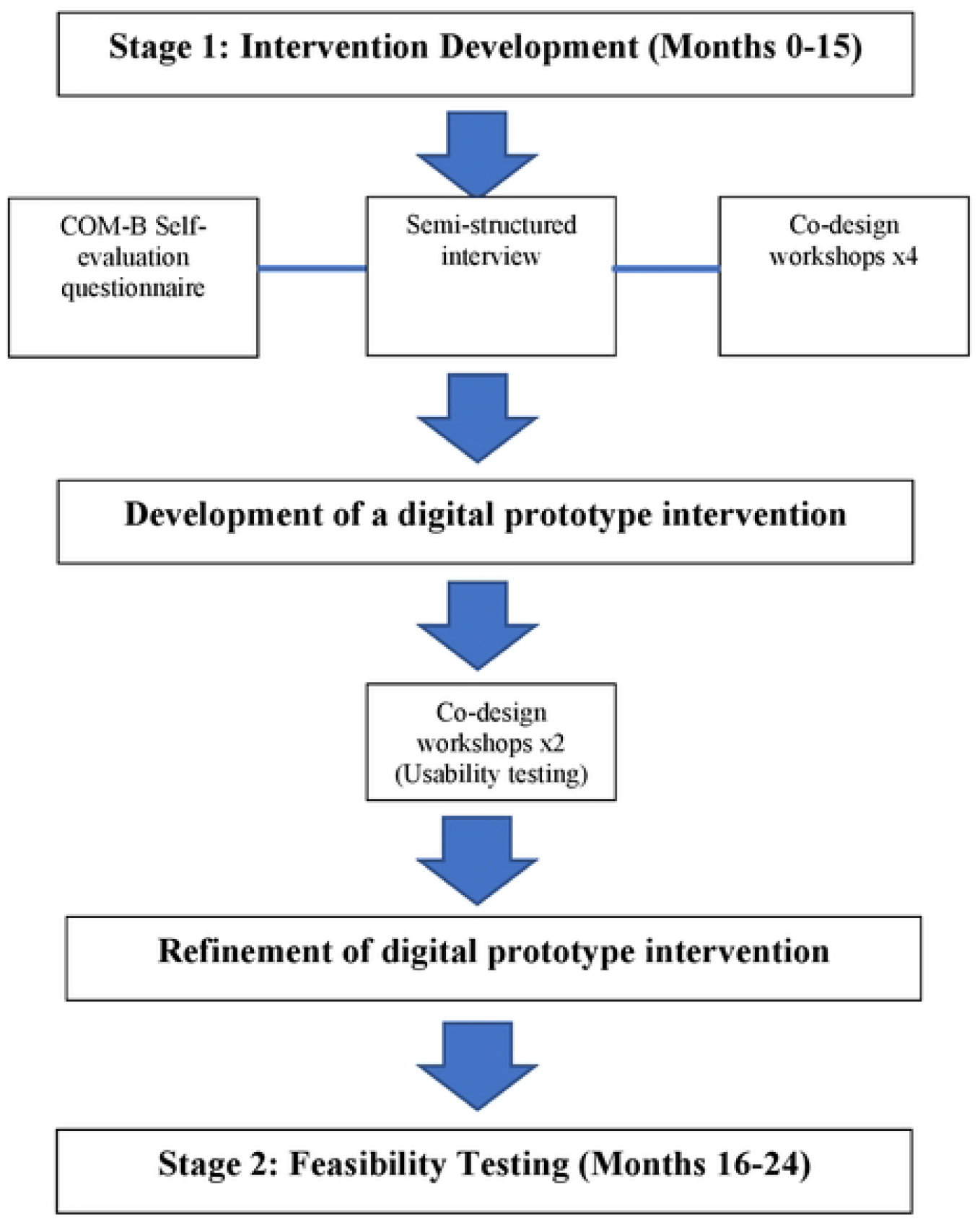
Overview of the study design and timelines.

**Figure 2:**
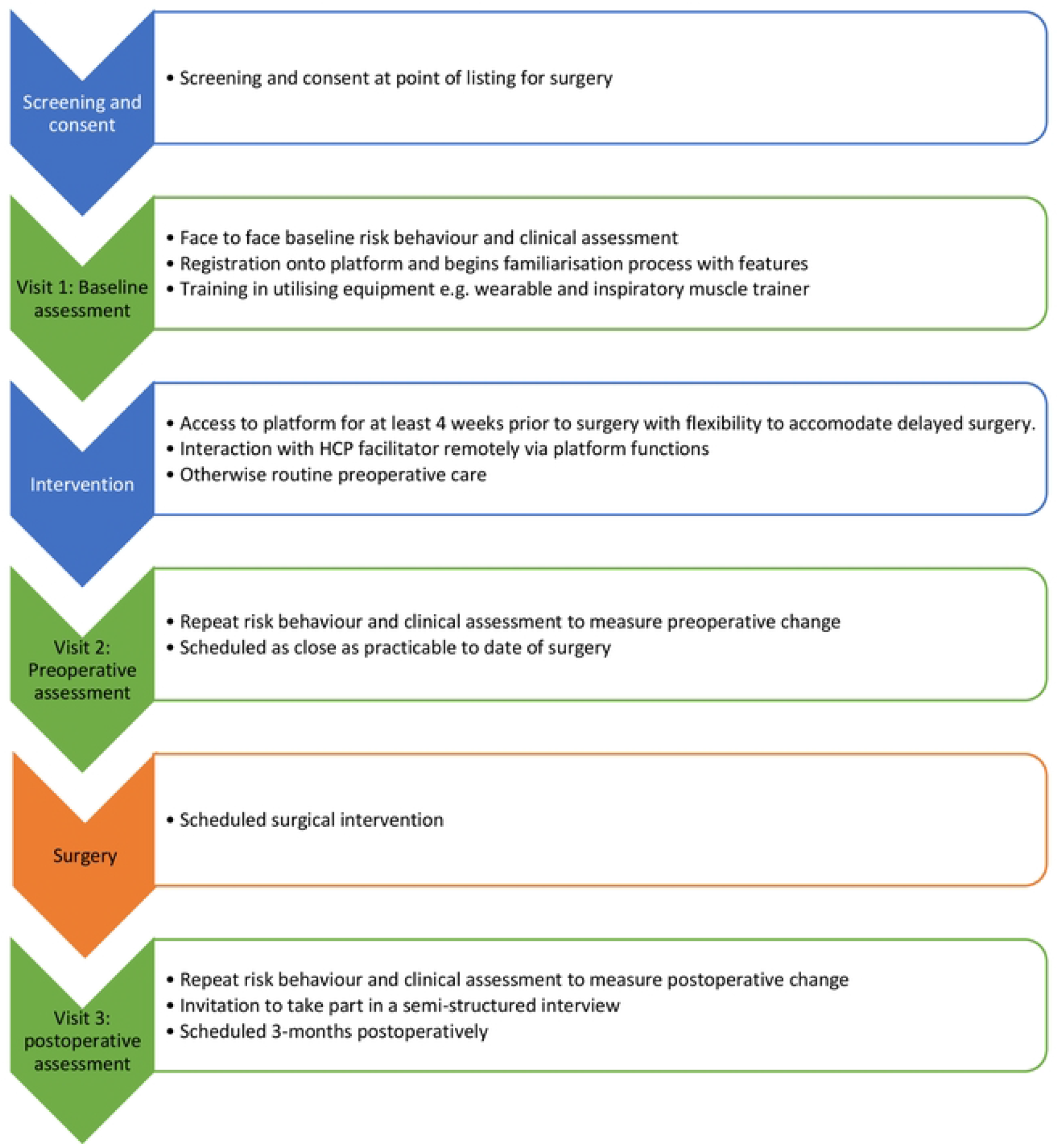
Overview of stage 2 of the study (feasibility study)

### Procedure

#### Stage 1 (Months 0-15)

A mixed method systematic intervention development process will be undertaken with reference to guidance for developing digital interventions [30]. The intervention will be underpinned and informed by the behaviour change wheel (BCW) [31], COM-B model, the theoretical domains framework (TDF) [32] and a person-based approach [33]. Data generated will inform the development of a logic model and selection of behaviour change techniques (BCTs) [34] for inclusion in the intervention. Subsequently, a co-design group will be recruited to collaborate with our multidisciplinary research team, including health psychologists, perioperative clinicians, exercise scientists, dietitians, and our partner web developers (Hark 2 Ltd, Leicester, UK). The group will include patient participants (those preparing for and having recently undergone major surgery), HCP participants recruited from the two participating NHS Trusts, and other stakeholders (e.g., commissioners) in order to develop a set of implementation strategies alongside the intervention [35].

##### Intervention

The multibehavioural digital intervention will be web-based and accessible via desktop, tablet, and mobile phone. The digital intervention and an accompanying training resource for HCPs will be co-designed with participants to facilitate changes in risk behaviours (e.g., physical activity, smoking, alcohol consumption, nutrition, sleep, and psychological wellbeing) with the overall aim to improve preoperative physical and mental health and wellbeing and reduce perioperative risk. The intervention will be designed for delivery/receipt over 4-8 weeks prior to surgery.

##### Stage 2 Overview (Months 16-24)

A single-arm mixed methods study will be used to assess feasibility and acceptability of the intervention with patients preparing for major surgery and HCPs promoting and supporting delivery of it each participating NHS Trust.

#### Stage 1 Sampling, eligibility criteria and recruitment

##### Sampling strategy

A purposive sampling strategy will be used to recruit patient and HCP participants representative of the UK major surgical population and the modern multidisciplinary perioperative team. In terms of patient recruitment, the aim will be to ensure maximal variation of age, gender, ethnicity, socioeconomic deprivation, and experience/confidence with online technology. Furthermore, we will aim to obtain a representative sample in terms of the health risk behaviours targeted by the prehabilitation intervention (e.g., smoking status). For recruitment of healthcare professionals, we aim to achieve maximal variation in terms age, gender, ethnicity, professional background, number of years in the role and experience with provision of prehabilitation support and digital healthcare interventions. Participant numbers recruited at each site will be adjusted to reflect differing surgical caseloads and specialties.

Up to 40 participants (20 patients and 20 HCPs) will be recruited to stage 1 of the study and asked to complete a COM-B self-evaluation questionnaire and participate in a semi-structured interview. With reference to published guidance on data saturation for theory-informed interview studies [36], an interim analysis will be conducted following data collection from the 10^th^ patient and the 10^th^ HCP participants. If new ideas and themes continue to emerge, recruitment will continue, and sample size will be increased in increments of three. This will be followed by a further interim analysis, up to a maximum of 20 patients and 20 HCPs. Where possible, these participants will also be invited to participate in co-design workshops.

Recruitment of participants to co-design workshops will be guided by individual session requirements. The aim is for patient and HCP participants to attend workshops together, with no more than 12 participants present at each session. Patient or HCP specific sessions may be required depending on progress of the co-design process and/or preferences of each participant group.

##### Eligibility criteria

###### Patients

Patients aged ≥18 years preparing for major surgery (as indicated by NICE NG45 [37]) or within 3-months of having undergone major surgery; discharged to their own home; able to communicate in spoken and written English, and able to provide informed written consent will be eligible to take part in the study. Patients receiving end-of-life-care will be excluded.

###### Healthcare professionals

Perioperative team members employed by participating Trusts from a medical, nursing, or allied healthcare professional background or a wider stakeholder in perioperative care (e.g., an individual with management or commissioning responsibility for perioperative services) will be eligible to take part. A willingness to take part in training to support promotion and/or delivery of the intervention is essential.

##### Recruitment and consent

###### Patient participants

Eligible patients will be identified by screening preoperative clinical and surgical lists by perioperative teams at participating Trust sites. A patient participant information sheet (PIS) will be sent by post or email to each participant, with a follow-up call within seven days to confirm receipt and determine interest in participation. Those wishing to take part in the study will be asked to provide informed consent prior to data collection through completion of a study consent form. Patients declining participation will continue to receive usual perioperative care and a reason for non-participation will be recorded.

We anticipate that patient participants may wish to involve a partner, friend, or family member during their interview or at workshops, and we acknowledge the valuable contribution these companions can make to the co-design process. As such, we will ask companions to complete a consent form to enable their contributions to be recorded, analysed and findings used to contribute to the intervention development process.

Preoperative patients and patients within 3 months postoperatively are eligible to participate in the study to inform intervention development. This acknowledges that short preoperative timeframes may prevent patients participating in all stage 1 components before their operation (e.g., major cancer surgery). Patients who do undergo surgery following participation in stage 1 of the study may continue to participate postoperatively if they wish. This facilitates the collation of views from patients who are approaching surgery and/or have undergone surgical intervention.

###### HCP participants

Eligible HCPs will be identified by clinical members of the study team and provided with a copy of the stage 1 HCP PIS by email. HCPs wishing to participate in the intervention development study will be asked to respond positively to the email invitation and subsequently provide informed written consent with a member of the research team prior to data collection. Additional recruitment will be undertaken to offset drop-out between stage 1 components.

#### Stage 1 Study procedures and data collection

A case record form will be completed for all stage 1 participants to facilitate a description of individual participants and to characterise the group overall. Baseline data to be collected from participating patients are demographics (i.e., age, sex, ethnicity, marital status, postcode for calculation of Index of Multiple Deprivations, and educational attainment); clinical and health risk behaviours (e.g., Surgical stage [pre/postoperative], surgical date/planned date, specialty and procedure/planned procedure, cancer status, Neoadjuvant chemoradiotherapy, comorbidities, Physical activity status [WHO criteria for healthy adults], smoking status, and alcohol intake [units per week]), malnutrition status [PG-SGA]; and information technology access and confidence (e.g., Frequency and availability of internet access, device ownership and utilisation). Baseline data to be collected from participating HCPs are, demographics (e.g., age, sex and ethnicity); and occupational data (e.g., clinical role, length of time in clinical role, prior experience in prehabilitation support, prior experience in utilisation of digital clinical interventions with patients).

##### COM-B self-evaluation questionnaires

The COM-B behavioural self-evaluation questionnaire adapted for the content of prehabilitation [31] will be administered to perform a behavioural analysis with each participant (patients and HCPs). In the context of behavioural change, capability (C), opportunity (O) and motivation (M) will be explored in accordance with the COM-B model. COM-B self-evaluation questionnaires are provided in our supplementary document (S1). Questionnaire data will be collated and used to inform and tailor semi-structured interviews.

##### Semi-structured interviews

Following questionnaire completion, participants will be invited to take part in a semi-structured interview with a research team member lasting up to 60 minutes. Interview topic guides [see supplementary document S2] will be informed by the COM-B model [31] and individualised to explore COM-B questionnaire responses in more detail.

##### Co-design workshops

A series of co-design workshops will be undertaken and facilitated by at least two members of the multidisciplinary research and design team. Each workshop will be guided by a schedule and will last up to two hours. The first workshop will involve a summary of the initial programme concept and COM-B questionnaire and semi-structured interview findings to provide context. Subsequent workshops will begin with a brief introduction, including session aims and objectives and progress made since previous workshops. Where workshops are conducted in-person, they will be conducted in line with each current covid-19 guidelines within each site to maintain staff and patient safety. Remote participation sessions (utilising a videoconferencing platform) will be offered if required (appropriate for ongoing pandemic restrictions). Given the nature of the intervention to be developed (i.e., remote/digital), it is considered appropriate to offer a remote option to participate to overcome barriers including cost and travel. Workshops will be supported by detailed notetaking by session facilitators.

Individual co-design workshops will be structured in response to findings from TDF analyses (see stage 1 data analysis) and activity during earlier sessions. Briefly, workshop topics will be informed by the findings of the behavioural analysis and TDF semi-structured interview findings with reference to the BCW and BCT Taxonomy v1 [34]. Participants will be invited to attend up to six workshops with no minimum commitment beyond one workshop. Workshops five and six will involve usability testing employing ‘think-aloud’ techniques [33]. The digital intervention content and associated HCP training intervention will be iteratively developed in collaboration with participants during each session. Following the conduct of the final workshop, the resulting prototypes will be updated/modified, where required in preparation for feasibility testing (i.e., stage 2 of the research).

#### Stage 1 Data analysis

##### Semi-structured interviews

All interviews will be audio recorded and transcribed verbatim. Transcripts will be thematically analysed (deductively) using the TDF. The following procedure will be followed: The first participant transcript will be independently pilot-coded by two team members and discussed to agree on an initial coding strategy. The same research team members will independently read, re-read and code two further transcripts. If a good level of agreement is achieved, the first researcher will code/analyse the remaining transcripts.

Text segments will be assigned to relevant domains of the TDF, and a thematic analysis conducted within each theoretical domain. If specific text segments do not fall into a specific TDF domain, additional domains will be generated to ensure the entire dataset is represented. Following analyses of the dataset, domains identified, and associated themes will be used to select BCTs to include within the intervention with reference to the Behaviour Change Taxonomy v1 [34].

##### Co-design workshops

Audio recordings of workshops will be transcribed verbatim. Transcripts will be reviewed alongside facilitator notes to capture all key information and decisions. This will enable an audit trail and reporting of when, how, and why key development decisions were made. Following the conduct of each co-design workshop, a summary document will be prepared to enable Hark 2 to iteratively develop an intervention prototype ahead of usability testing.

#### Stage 2 Feasibility study -Sampling, eligibility criteria and recruitment

##### Sampling strategy

Up to 40 patient participants listed for major surgery (from a range of surgical specialties) will be recruited to take part in the study from the two participating Trusts. This target sample size is informed by published guidance for pilot and feasibility studies [38] and accounts for potential drop-out (∼ 20%).

HCP participants will be recruited from each site and required to undergo training (training co-designed during stage 1) and either promote use of the digital intervention by patients or provide support to those using it. The number of stage 2 HCP participants will be guided by stage 1 findings (i.e., following consensus on who should fulfil what role).

##### Eligibility criteria

###### Patient participants

Patients aged ≥18 years preparing for major surgery (as indicated by NICE CG45 [36]) and available for a minimum of 4 weeks prior to planned surgery; ASA (American Society of Anaesthesiology) fitness for surgery ≥ grade 2; At least one health risk behaviour amenable to prehabilitation (e.g., current smoker); able to access and utilise the internet at home; able to communicate in spoken and written English, and able to provide informed written consent will be eligible to take part in the study. Participants who are pregnant or planning pregnancy; have severe mental illness (under active investigation or treatment by mental health services and/or preventing written informed consent); already undergoing prehabilitation or have a preference for an alternative mode of support (e.g., an in-person, face-to-face service); and those receiving end-of-life-care will be excluded. Where a patient participant has a safety contraindication to unsupervised exercise training based on ACSM criteria for clinical exercise testing and prescription [39], they will be excluded from the structured exercise component of the intervention but will be given access to other components of the intervention.

###### Healthcare professional participants

Perioperative team members currently caring for patients approaching major non-cardiac surgical intervention will be eligible to take part. A willingness to take part in training to support promotion and/or delivery of the intervention is essential.

##### Recruitment and consent

###### Patient participants

Patients listed for major surgery will be screened for eligibility by perioperative teams utilising electronic hospital records. Potential participants will be approached by telephone to explore interest. Those interested will be given a patient PIS sent by post or email. Interested patients will receive a follow-up telephone call by a team member within 7 days allowing time to receive, read and understand the study information and consider participation. Those who would like to participate will be invited to undertake a screening and baseline assessment (visit 1) where they will be given an opportunity to ask questions and complete a consent form with a study team member. Patients who decline participation at that stage will undergo routine preoperative care and their reason for non-participation will be recorded if they elect to provide one.

###### HCP participants

Perioperative team members at each site will be contacted by email inviting them to take part in the study with a follow-up after 7 days providing time to consider participation. The email will provide a HCP PIS and those who are interested in taking part will complete a consent form with a study team member and be invited to begin the intervention HCP training package.

### Stage 2 Outcome measures

#### Primary outcomes

##### 1. Feasibility

Feasibility will be determined by assessing participant recruitment and retention rates, time taken to recruit to the target sample size, and rates of intervention uptake and completion, including number of patients completing all relevant components of the intervention.

Feasibility of the training intervention will be determined by assessing HCP participant recruitment and retention rates, time taken to recruit to the target sample size, and rates of training intervention uptake and completion, including willingness to refer to the intervention and continue to promote and support patient participants with the intervention.

##### 2. Fidelity

Fidelity of delivery will be assessed by collecting data relating to intervention usage by patient participants, including when components were accessed, revisited, and for what length of time. Fidelity of receipt and enactment will be assessed qualitatively via semi-structured interviews with patient participants.

Fidelity of delivery of the training intervention will be assessed by audio recording delivery of the training to ensure all intervention components are delivered per protocol using an intervention fidelity checklist [40]. Fidelity of receipt and enactment will be assessed qualitatively via semi-structured interviews with HCP participants.

##### 3. Acceptability

Acceptability will be assessed quantitatively and qualitatively. In terms of patient participants, data will be collected on the number of logins over the intervention period and the number of interactions with facilitating HCP participants. In terms of HCP participants, data will be collected on the number of HCPs who consent to take part in the study/be trained and who complete training. Semi-structured interviews using the TDF as an analysis framework will obtain participant (patients and HCPs) views and experiences of the intervention, including their experiences of using/interacting with the intervention, perceived barriers, and facilitators to using it and suggestions for ways in which it could be improved.

### Stage 2 secondary outcomes

Data will be collected on the following secondary outcomes: Patient activation (Patient Activation Measure [PAM]); physical activity (International Physical Activity Questionnaire [IPAQ], accelerometery data from integrated wearable device); smoking status (self-reported); alcohol consumption (units per week); nutritional (PG-SGA) and dietary status (Dana-Faber healthy eating questionnaire, modified for personal consumption); sleep (Pittsburgh Sleep Quality Index); exercise capacity (6-minute walk test [6MWT], 30-second sit to stand repetitions, grip strength, maximum inspiratory pressure); Psychological wellbeing (Hospital Anxiety and Depression Scale [HADS]); Health-related quality of life (HRQOL using SF-36v2 and EQ-5D-5L); postoperative mortality and morbidity (30 and 90 day mortality, Comprehensive Complication Index [CCI]); and length of stay and readmission (length of hospital stay, length of critical care stay, days at home [or usual residence] within 30 days of surgery [DAH_30_]). The feasibility and sensitivity of data collection for these outcome measures will be explored to identify candidate primary outcome measures for a future randomised controlled trial of the intervention.

In addition, semi-structured interviews will qualitatively assess feasibility and usability of the integrated wearable device in support of programme components and perioperative biometric monitoring.

#### The digital intervention (iPREPWELL)

The content and format of the digital intervention components will be informed by the systematic development process undertaken during stage 1 of the study. However, the intervention will have the following features and functions:

1. Intervention duration – the time between participants being listed and having their surgery is between 4 and 8 weeks on average, therefore the duration of the intervention will run in accordance with this timeline. Access will be continuous during this time and up to 3 months postoperatively.
2. Intervention components offered to participants will be personalised during registration, i.e.., non-smokers will not be offered content related to smoking.
3. Given the tendency for clustering of health risk behaviours and limited preoperative timeframes in surgical populations, intervention components will be designed to run simultaneously. They will be delivered using textual, audio, and visual material. Decisions about the specific mode of delivery and format of each intervention component will be informed by findings from the systematic development process.

Additional intervention features could include:

- Incorporation of a wearable physical activity monitoring device to facilitate self-monitoring and real-time participant feedback. The most appropriate device will be agreed in collaboration with participants during phase 1 of the study.
- An online forum facilitating interaction with facilitators and other participants.
- Direct messaging between the facilitating HCP and participants to prompt behavioural change and provide support.
- Access to educational content in the context of the perioperative journey (e.g., ‘digital surgery school’).

The physical activity and exercise component of the intervention will be included for all participants reflecting the high rates of physical inactivity within this clinical population, and the potential to enhance aspects of physical fitness in surgical populations [11]. Only participants with identified contraindications to physical activity or exercise will be excluded from this component of the intervention. This intervention component will support increased physical activity and remotely supervised structured exercise before surgery including aerobic, resistance/strength and inspiratory muscle training. Specifically, this will include:

- Provision and use of home-based exercise equipment, including resistance bands and an inspiratory muscle training device
- Utilisation of the integrated wearable device to guide training sessions and provide feedback e.g., heart-rate guidance for aerobic training sessions

Patients will be encouraged to login throughout the intervention period to engage with the various components to promote/maintain motivation and volition to support health behaviour change. It is anticipated that patients will require a level of remote HCP support throughout the timeline of the intervention. What this involves will be determined during stage 1 of the study, the developmental process. HCP participants will take part in training prior to supporting patient participants.

#### The training intervention

The content and format of the training intervention for HCPs will be informed by the systematic development process undertaken during stage 1 of the study. Not wishing to pre-empt the outcome of stage 1 of the study, training is likely to incorporate health behaviour-specific content to target knowledge, and skills-based training to facilitate promotion of the intervention during routine care and to facilitate the provision of support to patients throughout the intervention period. The training intervention, as with the patient intervention, will be theory and evidence-informed with reference to the BCW [31].

#### Study visits

Figure. <u>3</u>2 provides an overview of stage 2 of the study (feasibility study).

#### Visit 1 (Screening and baseline assessment)

Patient participants will attend the hospital site to undergo a baseline assessment process (incorporating a safety screen for remotely supervised exercise based on ACSM guidance [38]) and registration onto the intervention. The assessment will combine clinical, health behaviour and exercise capacity elements as presented earlier. It will be conducted by a facilitating HCP participant and at least one research team member. The methods for physical activity and exercise capacity assessments are provided in our supplementary document (S3). Following visit 1, patient participants will utilise the digital intervention at home with remote support by a trained HCP participant.

#### Visit 2 (preoperative assessment)

Visit 2 will be scheduled prior to surgery to assess changes in health behaviours (e.g., physical activity) following platform usage. The visit will be conducted at the hospital site by at least two research team members. Data collected will mirror visit 1 (supplementary document [S4]).

#### Visit 3 (postoperative assessment)

Visit 3 will be scheduled at 30 days postoperatively to assess change/maintenance of health behaviours and to collect postoperative outcome data. The visit will be conducted at the hospital site by at least two research team members. Data collected will mirror visits 1 and 2.

Figure. 3 presents the stage 2 schedule of enrolment interventions and assessments at each study visit timepoint based on SPIRIT recommendations for clinical trials.

**Figure 3.**
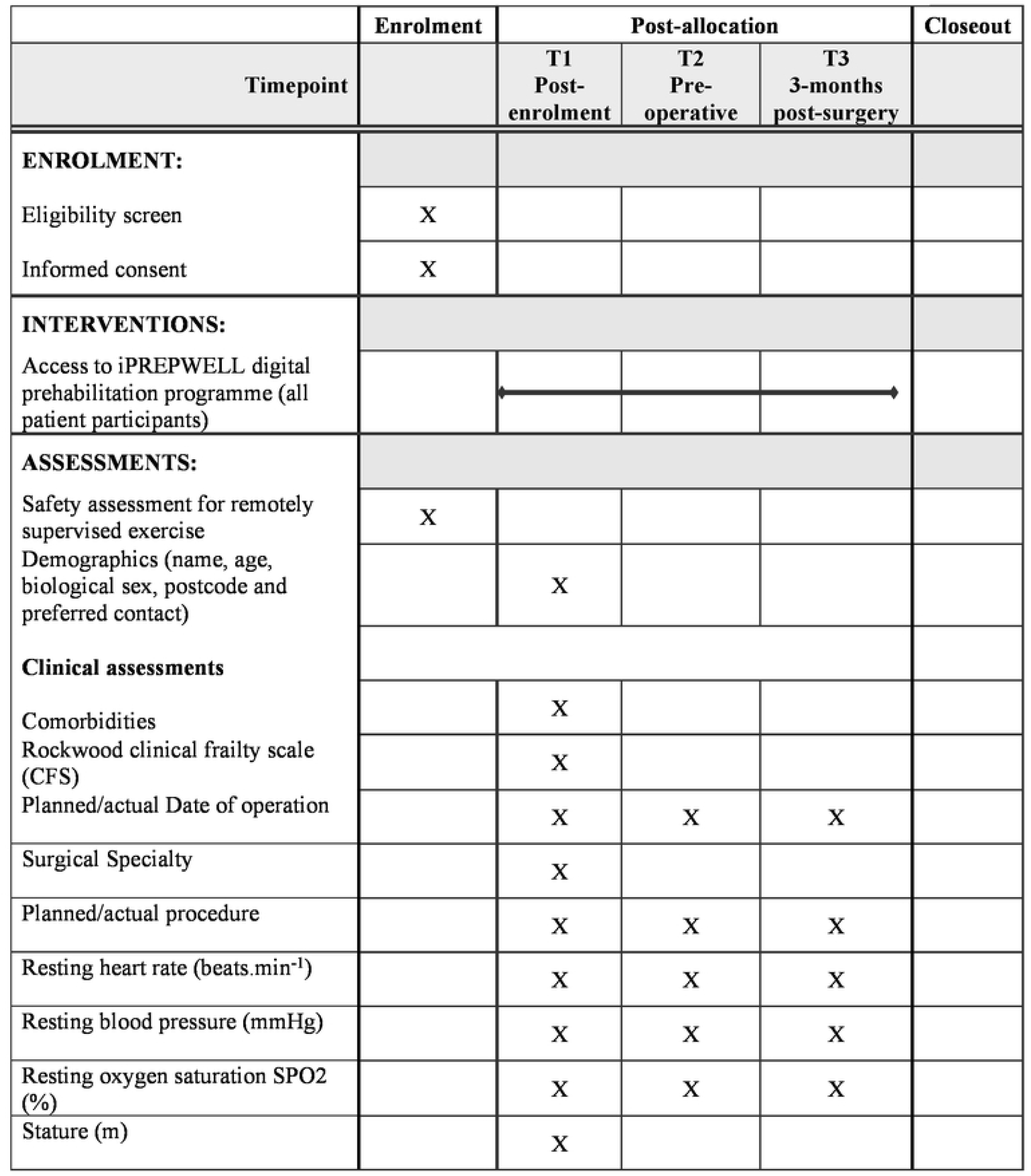
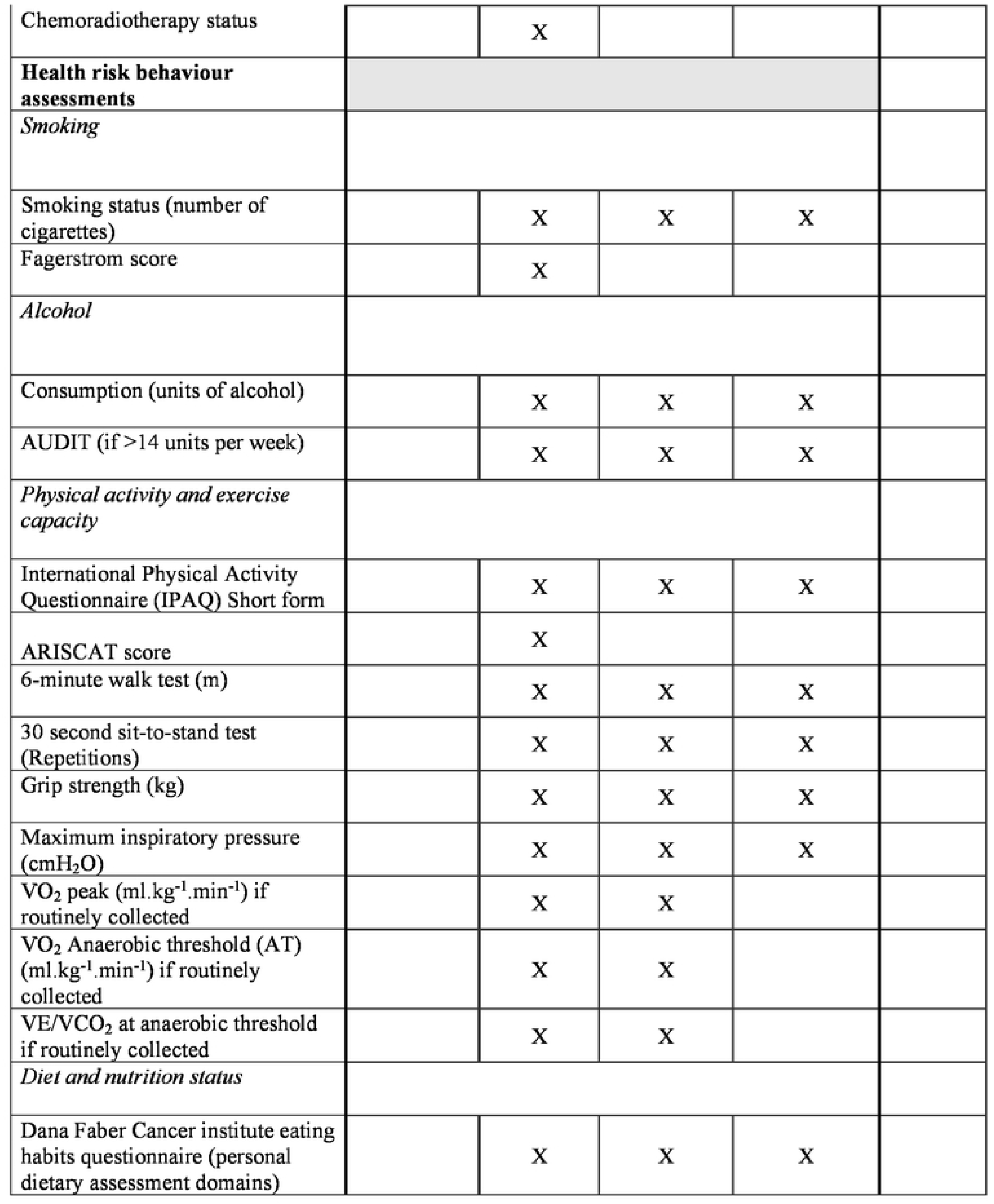
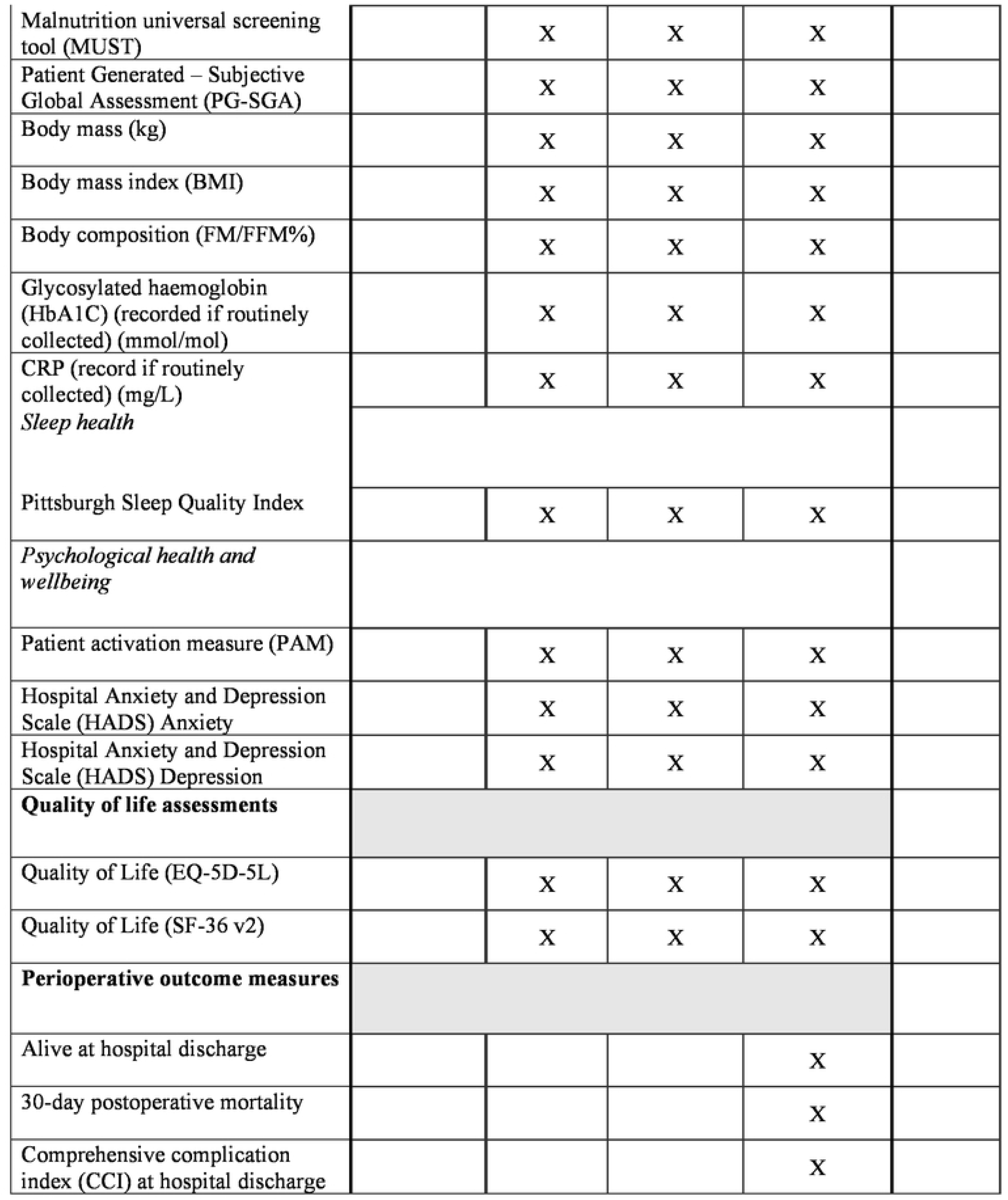
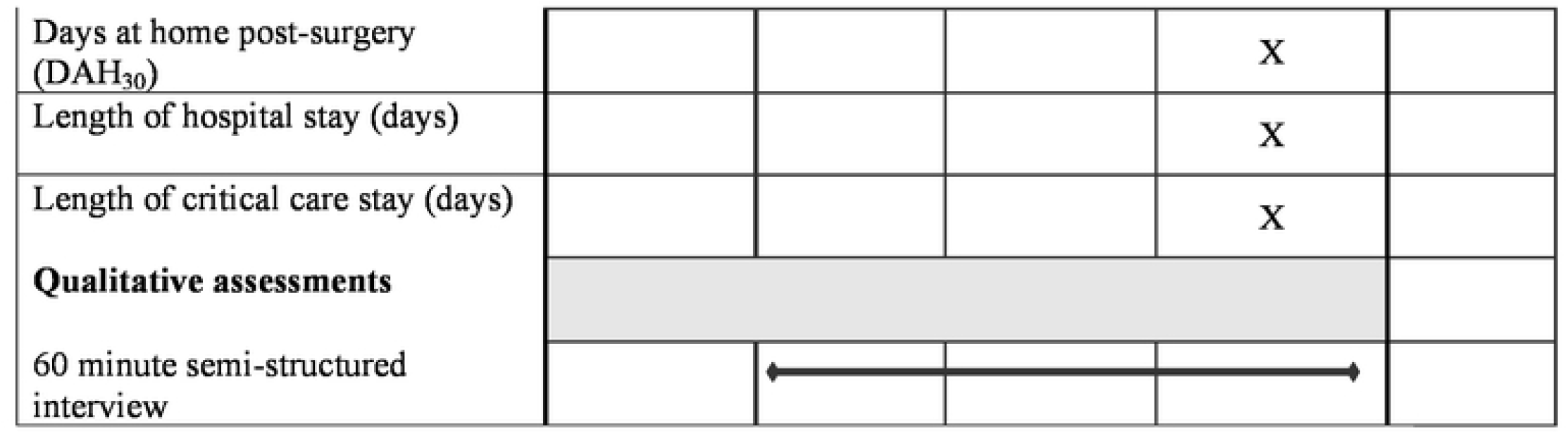
**Study stage 2 SPIRIT schedule of enrolment, interventions, and assessments**.

#### Quantitative data collection

Data will be collected, where possible, as an electronic case-record form (e-CRF) within the online intervention. Completion of these will be scheduled as part of intervention utilisation e.g., the registration process will include e-CRF 1. Data will be entered by patient participants, with additional data input by HCP participants and study team members, where appropriate.

Additional data will be collected on intervention utilisation, e.g., number of logins, duration of session, completion of individual intervention components, and information entered by participants during intervention usage. The integrated physical activity wearable device will collect data that will be uploaded into the intervention platform, stored, and made available to participants, e.g., daily recorded step counts.

#### Qualitative data collection

Up to 40 patient participants and all participating HCPs will be invited to take part in a semi-structured interview with a research team member. This component of the study is optional (i.e., patient participants can take part in the intervention study and refuse participation in the qualitative study). In keeping with stage 1, companions will also be included if patient participants wish and will complete a stage 2 consent form to allow their interview contributions to be included in the analysis. All interviews will be audio recorded and transcribed verbatim.

To facilitate an early health economic analysis, HCP participants will be asked to complete a diary of activity in terms of support provided to patient participants.

### Stage 2 Data analysis

#### Quantitative data

Data will be summarised descriptively using mean and SD or median and IQR for continuous variables, and count and percentage for categorical variables. As this is a feasibility study, the level of missing data will be documented but no imputation undertaken.

An initial health economic analysis will be conducted to focus on costs of intervention delivery to inform design of a future efficacy study.

An initial exploratory analysis of pseudo anonymised perioperative wearable data will be undertaken utilising machine learning techniques supported by Telstra Health UK.

#### Qualitative data

Qualitative data will be thematically analysed using the TDF. Two members of the research team will independently code and analyse interview transcripts. The same procedure will be undertaken as described during stage 1 to develop a coding strategy.

A detailed description of how data will be handled is provided in supplementary document (S4).

#### Study management

A study management group (SMG) will be established by the chief investigators prior to the commencement of stage 2 of the study with representation from the sponsor, participating sites and institutions, patient representatives recruited during stage 1, and research partners. The group will oversee the conduct of the feasibility study and meet monthly, or as required.

#### Study Safety considerations

##### Stage 1

Participation during Stage 1 is anticipated to present a low risk of adverse events (AEs) for participants. Potential AEs occurring during stage 1 activities will be assessed, graded, and followed up until resolution by the study team in keeping with study sponsor and UK Good Clinical Practice (GCP) guidance.

##### Stage 2

Potential AEs occurring throughout the duration of stage 2 of the study, whilst the intervention will be assessed, graded, and followed up by the research team until resolution in keeping with sponsor and GCP guidance.

Risk to patient participants is most likely to originate from participation in the structured exercise training programme. Other intervention components are not anticipated to lead to AEs. The overall risk of AEs relating to exercise is considered low. This is based on a growing body of evidence demonstrating the safety of structured exercise training (including aerobic, resistance and inspiratory muscle training) in surgical populations [41]. This is in addition to the safety profile of several hundred maximal effort cardiopulmonary exercise tests conducted in the study target population at participating sites and nationally [42].

Despite this, we are mindful of the additional risk this poses in comparison to directly supervised exercise interventions. The following measures are planned to mitigate this as far as possible:

- An independent clinician will review all serious adverse events (SAEs) and report to the study management group.
- Participants will be formally risk assessed to confirm safety for participation based on international criteria for exercise training and testing [42] and the expertise of an active face-to-face surgical prehabilitation service.
- Participants will undergo several functional capacity assessments face-to-face with trained healthcare professionals prior to commencing remotely supervised training.
- The exercise intervention will begin with clear, co-designed safety instructions relating to both undertaking physical activity safely and undertaking activity outside the home environment.
- Clear channels for participants will be provided to raise non-emergency concerns with HCP facilitators and the research team and how to access help in an emergency.
- The exercise component of the intervention will be scaled to participant capabilities and progression in intensity will be participant, rather than facilitator lead.
- Wearable data collected during training sessions will allow intensity monitoring and adjustment as required.

#### Stage 2 participant discontinuation and withdrawal

Stage 2 participants will be free to withdraw from the study at any stage without providing a reason.

Participant discontinuation will occur with any of the following:

- Completion of the stage 2 study protocol.
- Acute Illness requiring hospital admission
- Death of participant or commencement of end-of-life care
- Decision to cancel surgical intervention
- Loss of capacity to consent to continue participation
- Participant decision to withdraw
- Investigator decision
- Study management group or chief investigator decision
- Severe non-compliance to protocol as judged by the investigator and/or sponsor
- Safety reasons

If a participant wishes to withdraw or is discontinued from the study, the following procedures will be observed:

- Participants will be offered the chance to take part in a semi-structured interview to provide their reasons for withdrawal from the process to allow learning. Participants will be free to decline this interview without providing a reason.
- Withdrawal of consent/ discontinuation of the study will be clearly documented in study documentation and the participant’s medical record.
- No further clinical data will be collected from the participant. However, existing clinical data held will be retained and used for the research.
- Patients will continue with standard of care treatment as recommended by their treating team.

### Approvals and registrations

Ethical and regulatory approval for the study has been obtained from Health Research Authority (HRA) North West Preston Research Ethics Committee (Ref: 21/NW/0219). The study is registered on the ISRCTN registry (ISRCTN 17788295) and has been adopted onto the UK National Institute for Health and Care Research (NIHR) portfolio for anaesthesia, pain, and perioperative medicine with South Tees Hospitals NHS Foundation Trust as study sponsor (contact details available via corresponding author).

#### Study status and timeline

Stage 1 study recruitment is underway at time of writing and commenced in October 2021. The study is planned to complete by October 2023.

## Discussion

We have presented a protocol for the development and feasibility testing of a theory-informed co-designed, multibehavioural prehabilitation intervention for people preparing for major surgery. At the time of writing, we are unaware of any robust developed interventions following a systematic developmental process available to target changes in multiple health behaviours simultaneously, which is an urgent unmet need in perioperative care. This study aims to develop, and feasibility test a digital multibehavioural intervention for patients and a training intervention for healthcare professionals.

We acknowledge several important limitations to the protocol for the study at this stage. Firstly, our study will be conducted at two centres in the North of England (UK) which may limit wider applicability. Although, both centres serve geographically and socioeconomically diverse populations that will offset this to some degree and this will be further mitigated by a purposive sampling strategy to ensure maximum variation in stage 1 participants. Secondly, we will develop an intervention for those approaching major surgery. We acknowledge this may result in an intervention that is not fully optimised for specific surgical populations or pathways. However, this is deliberate to produce a generic intervention that is feasible and acceptable for the majority of surgical patients and can be readily modified and adapted for specific populations going forward. Should the intervention developed demonstrate to be acceptable and feasible by participating patients and HCPs, a further study will be required to establish effectiveness and cost-effectiveness. Finally, the absence of a control arm within the feasibility study for reasons of time-efficiency and study cost will prevent assessment of intervention efficacy. However, this is not the main aim of the study and the data collected with the single-arm design will provide useful data in support of any follow-up efficacy trial.

Stage 1 and stage 2 findings of this study are planned to be disseminated by peer-reviewed publication and presentation at relevant conferences. In addition, our wider study team have links to regional and national initiatives to improve the readiness of patients approaching major surgery in the wake of the Covid-19 pandemic, offering broader opportunities to evaluate and scale the developed programme if the findings of this study support this.

Study amendments will be by submission to the approving Research ethics committee in accordance with UK HRA policies and procedures. Study termination will be either planned by completion of the full protocol at both participating sites or unplanned by the chief investigators following consultation with the study management group.

## Data Availability

o datasets were generated or analysed during the current study. All relevant data from this study will be made available upon study completion.

N.A

## Authors’ contributions

**Conceptualisation:** JD, GD, LA, GT, DY

**Methodology:** All authors

**Formal analysis:** JD, RL, LA, GT, CG

**Writing** – original draft: JD, LA

**Writing -review and editing:** All authors

## Acknowledgements

The authors would like to offer thanks to Sport England, Macmillan Cancer Support and for their financial support of this study. The fundings bodies will not have involvement or authority in study design; data collection, study management, data analysis and interpretation of data; report writing or the decision to submit the report for publication.

The authors have no conflicts of interest to declare in the context of this study.

## Supporting information

**S1: Stage 1 Patient and HCP participant COM-B self-evaluation questionnaire**.

**S2: Stage 1 Patient and HCP participant semi-structured interview topic guides**.

**S3: Methods for stage 2 physical activity and exercise capacity assessment**

**S4: Stage 2 Data collection and methods at scheduled study visits**

**S5: Data Handing**

